# Directional interface mechanics using magnetic resonance elastography predicts focal tumor recurrence in glioblastomas

**DOI:** 10.64898/2026.05.06.26352294

**Authors:** Jan Saip Aunan-Diop, Ancuta Ioana Friismose, Ziying Yin, Emi Hojo, Sandeep Ganji, Yuan Le, Frederik Harbo, Bo Halle, Frantz Rom Poulsen

## Abstract

Glioblastoma progression is spatially heterogeneous, but conventional imaging provides limited information about where subsequent tumor progression is likely to occur. We developed a directional magnetic resonance elastography (MRE) framework to test whether local post-treatment tumor–brain interface mechanics are associated with later spatial tumor progression. In a secondary analysis of a prospectively acquired glioblastoma cohort, wedge-level viscoelastic instability features were extracted from the first post-treatment MRE scan and related to novel tumor burden on the second post-treatment scan after excluding tumor already present on pretreatment or first post-treatment imaging. Nine patients had longitudinal imaging suitable for spatial comparison; six lesions showed net interval growth and were included in the primary wedge-level directional analysis, while three non-growing lesions were retained for descriptive comparison. In growing lesions, several directional mechanical features were descriptively associated with later novel tumor burden. In cluster-aware models accounting for within-patient dependence among wedges, mean Δtanδ showed the most consistent association with later wedge-level novel tumor fraction across mixed-effects and generalized estimating equation analyses. Associations were directionally stable across wedge-width sensitivity analyses. These findings provide proof of principle that post-treatment glioblastoma interface mechanics contain spatially resolved information related to where later tumor emergence occurs, supporting further validation of directional MRE as a framework for longitudinal mapping of progression geometry.

## Introduction

Glioblastoma (GBM) progression is spatially heterogeneous, with biologically distinct tumor core and edge compartments, regionally variable treatment response, and disease evolution that changes across both time and space^1,2^. Even after treatment, tumor progression or regrowth rarely develops as uniform circumferential expansion. Instead, new tumor often emerges preferentially along selected parts of the lesion boundary, suggesting that progression is shaped by local tissue conditions at the tumor–brain interface rather than by lesion-wide state alone^3^. Conventional magnetic resonance imaging (MRI) is central to clinical follow-up, but it provides little information about where future tumor extension is likely to occur before overt structural change becomes visible^3–5^. A biomarker that identifies spatially vulnerable regions before overt progression would therefore be of substantial biological and clinical interest.

Mechanical interaction parameters are plausible candidates for such a biomarker^6–10^. Glioblastoma grows within a mechanically complex environment in which tumor cells, extracellular matrix, interstitial fluid, and surrounding brain tissue interact dynamically across the lesion boundary^11–15^. These interactions are unlikely to be spatially uniform. Instead, local differences in tissue coupling, stress redistribution, viscous dissipation, and structural coherence may create directional conditions that favor later extension^2,10,14,16–19^. In this framework, progression is not simply a function of how abnormal a tumor is overall, but of where along the interface mechanical disequilibrium is greatest.

Magnetic resonance elastography (MRE) provides non-invasive access to tissue viscoelastic properties in vivo and is therefore well suited to probe such processes^20,21^. Beyond stiffness alone, MRE resolves storage and loss-related mechanical behavior and can depending on the reconstruction model, can estimate parameters such as the loss tangent. These quantities reflect distinct aspects of tissue mechanics and may be altered by tumor-induced disruption of microstructure, fluid redistribution, necrosis, and local failure of tumor–host mechanical coherence^6^. Prior work has shown that glioma fluidity and viscoelastic state are biologically informative, and recent in vivo modeling has linked glioma-associated deformation and stiffness gradients to excess solid stress and unstable interface conditions^6,10,11,22–28^. Together, these observations support the broader idea that glioblastoma progression may be mechanically structured at the tumor boundary rather than isotropic in space.

Our recent work has further suggested that such structure may be detectable as a mechanically unstable interface phenotype^6,29^. In cross-sectional analysis, glioblastomas showed diffuse, branched, and fragmented instability fields compared with the more coherent and radially ordered patterns seen in meningiomas, consistent with viscoelastic decoupling at the tumor–brain interface^6^. In a separate post-radiotherapy study, normalized viscoelastic parameters provided the clearest inferential separation between radiation necrosis and recurrent metastasis, while exploratory interface-topology features contributed complementary spatial information^29^. These findings collectively suggest that MRE may be most informative when mechanics are treated as spatially organized rather than reduced to lesion-wide summary values.

What remains unclear is whether such local mechanical abnormalities have longitudinal spatial consequences. Existing MRE studies have largely focused on cross-sectional classification, lesion-level associations, or global treatment response^11^. Much less is known about whether local post-treatment mechanics anticipate the later geometry of tumor emergence. Addressing this question requires a framework that links mechanical features measured at one time point to spatially matched tumor progression at a later time point.

Here we developed a directional wedge-based longitudinal MRE framework to test whether local post-treatment interface mechanics are associated with subsequent novel tumor emergence in the same directional sector. The analysis was performed on the maximal tumor slice of the first post-treatment scan, where the peritumoral interface was subdivided into directional wedges and wedge-level mechanical features were extracted. Later novel tumor burden was then quantified in the matched wedges on the second post-treatment scan after excluding tumor already present on the pretreatment or first post-treatment scans. We hypothesized that sectors with more abnormal post-treatment mechanics would show greater subsequent novel tumor burden, consistent with a spatially structured biomechanical basis for glioblastoma progression.

## Methods

### Study design and cohort

We performed a secondary longitudinal imaging analysis using data from a prospectively acquired cohort of GBM patients with post-treatment MRE and tumor segmentations available at multiple time points. All subjects provided oral and written informed consent prior to inclusion and the study was approved by the Regional Committee on Health Research Ethics for the Region of Southern Denmark (IDs: S-20190105, S-20220055) and was performed according to the declaration of Helsinki. The primary analysis linked directional mechanical features measured at the first post-treatment scan to subsequent novel tumor burden measured at the second post-treatment scan. The working hypothesis was that local abnormalities in tumor–brain interface mechanics at the earlier scan would be associated with later tumor emergence in the same directional sector.

Patients were eligible for the longitudinal directional analysis if they had an analyzable first post-treatment MRE examination, tumor segmentation at the first post-treatment scan, and a second post-treatment MRI scan suitable for spatial comparison of interval tumor change. Cases were excluded if the first post-treatment MRE data, the first post-treatment tumor segmentation, or the second post-treatment follow-up MRI were unavailable or not suitable for analysis.

The primary inferential analysis was restricted to lesions with net interval growth between the first and second post-treatment scans, defined by the change in total segmented tumor volume across these time points. Lesions without net interval growth were retained for descriptive analyses only, because they did not provide a positive directional endpoint for modeling subsequent tumor emergence.

### Image acquisition, registration, and segmentation

MRI and MRE were performed on a 3T clinical system (Achieva, Philips, The Netherlands) using a 16-channel head coil^30^. The protocol included anatomical MRI and 60 Hz MRE acquired using a pillow-like passive driver positioned under the head. Wave propagation was imaged in a three-dimensional volume with motion sensitization in three orthogonal directions using a spin-echo echo-planar imaging sequence with eight phase offsets. Acquisition parameters were TR 4800 ms, TE 67 ms, FOV 240 × 240 mm^2^, acquisition matrix 80 × 80, slice thickness 3 mm, 48 contiguous axial slices, and parallel imaging acceleration factor 2. After acquisition, the MRE magnitude and phase images were post-processed using an offline custom script to compute MRE-derived maps, including stiffness, loss modulus, and storage modulus.

Registration and segmentation were performed in 3D Slicer^31^. All post-treatment images were rigidly co-registered to the pretreatment MRE magnitude image and corresponding parameter maps. Contrast-enhancing tumor regions and peritumoral T2/FLAIR hyperintensity were manually segmented on pretreatment images and propagated to the post-treatment scans with manual adjustment when required by a neuroradiologist (FH). In cases where contrast enhancement was reduced after treatment, the pretreatment contrast-enhancing contours were retained when persistent T2/FLAIR abnormality indicated tumor presence. Regions with necrosis were excluded. A normal-appearing white matter (NAWM) ROI was used for normalization. The resulting longitudinal tumor masks served as the geometric input for the directional wedge analysis.

### Directional wedge framework and outcome definition

The directional analysis was performed on the axial slice containing the largest segmented tumor area on the first post-treatment scan. This maximal tumor slice served as the geometric basis for directional sampling and for quantification of later novel tumor burden.

Within this slice, the tumor centroid was defined as the mean row–column coordinate of all tumor voxels. Directional sampling was performed over 72 evenly spaced angular directions, corresponding to 5-degree increments across 360 degrees. For each direction, a wedge was defined as an angular sector extending radially outward from the tumor boundary. A peritumoral shell was constructed outside the tumor boundary by including voxels within 6.0 mm of the boundary but external to the tumor mask, and excluding voxels outside the brain parenchyma. In-plane voxel spacing was estimated from the storage-modulus image and used to convert physical distance to pixel units. Directional wedges were obtained by intersecting this shell with angular sectors centered on each sampled direction.

The primary analysis used a wedge half-width of 10 degrees. Additional analyses were performed at wedge half-widths of 5 degrees and 15 degrees. Because adjacent wedges were centered on closely spaced directions, neighboring wedges overlapped partially, providing directional smoothing while preserving angular specificity. The workflow is presented in **Figure 1**.

**Figure 1:**
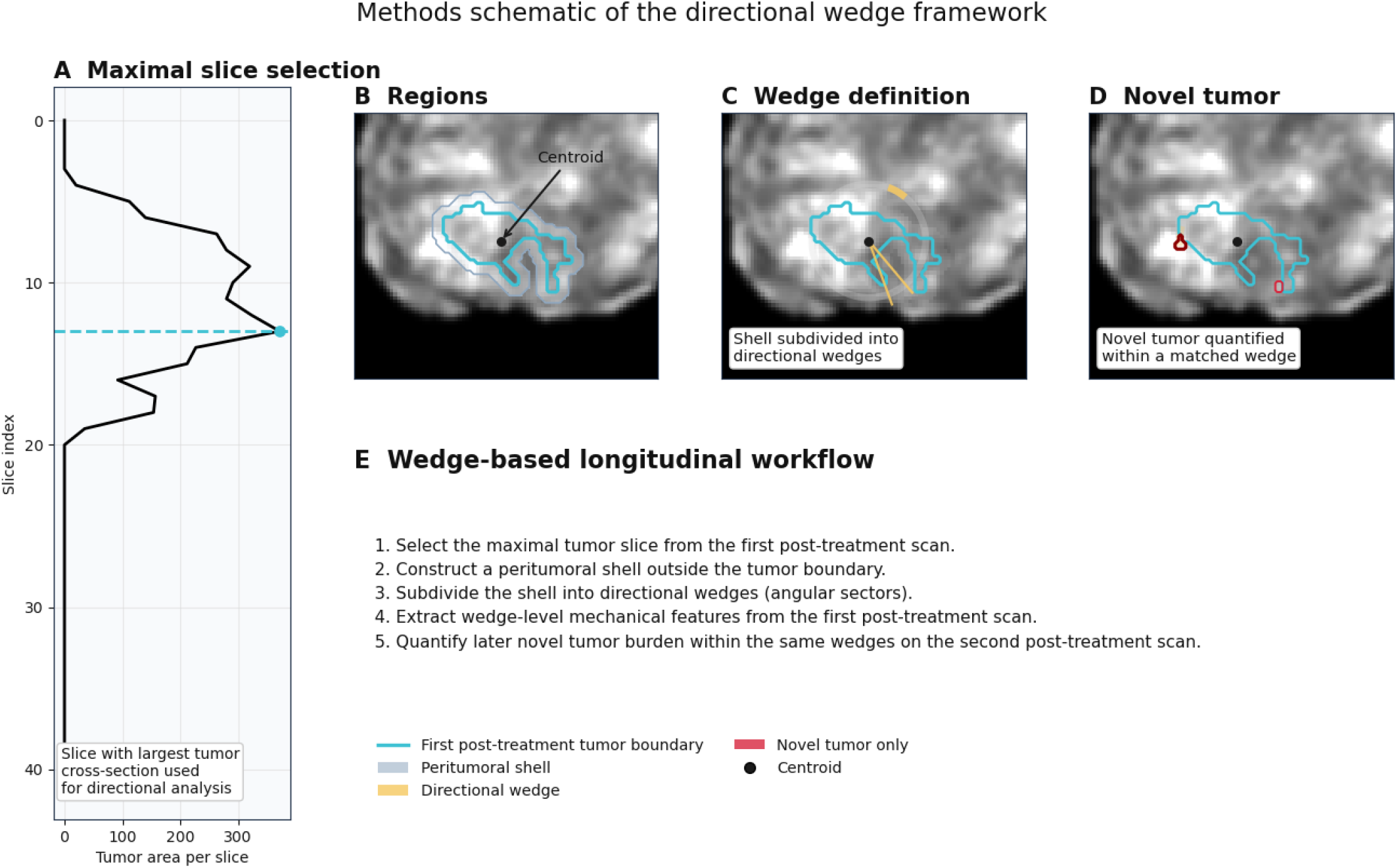
**A** The axial slice containing the largest tumor cross-section on the first post-treatment scan was selected for directional analysis. **B** A peritumoral shell was constructed outside the tumor boundary, and the tumor centroid was used as the angular origin. **C** The shell was subdivided into directional wedges, defined as angular sectors centered on evenly spaced directions. Wedge-level mechanical features were extracted from the first post-treatment scan, and later **D** novel tumor burden was quantified on the second post-treatment scan within matched wedges after excluding tumor already present on the pretreatment or first post-treatment scans. **E** Instability metrics were used to predict new tumor in wedges.

The primary outcome was wedge-level novel tumor burden (progression) on the second post-treatment scan. Novel tumor was defined conservatively as tumor present at the later post-treatment scan but absent from both the first post-treatment tumor mask and the pretreatment tumor mask. Within each wedge, the primary outcome was defined as the fraction of wedge voxels occupied by novel tumor at the later scan (novel_fraction_in_wedge). A secondary binary outcome indicated whether any novel tumor was present within the wedge (novel_present_in_wedge).

### Mechanical feature extraction

**T**he instability index has been described previously as a voxel-level mechanical measure intended to capture the combined effect of altered viscous behavior and altered phase relationship in tumor tissue^6^. Briefly, the original formulation was:

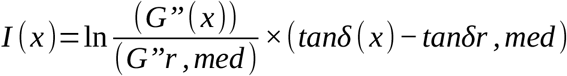

where *G*’’ (*x*) is the voxel-wise loss modulus, 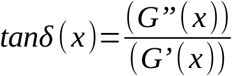 is the voxel-wise loss tangent, and *G’’r, med* and *tanδr, med* denote median reference values from the designated reference region. In the present study, the directional wedge framework required a local image-based implementation that could be applied consistently across all analyzed cases and wedge regions. Mechanical features were therefore extracted exclusively from the first post-treatment scan, and the instability-related quantities were implemented using image-wide median reference values across valid voxels within the same post-treatment image volume.

**S**torage modulus and loss modulus maps were used to derive three voxel-level fields. First, a log-transformed loss-modulus ratio was calculated as:

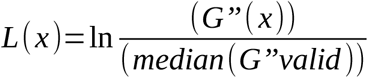

Second, the loss tangent was defined as:

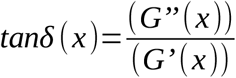

and its deviation from the image-wide median reference value was computed as:

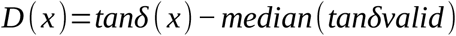

Third, a composite instability field was defined as:

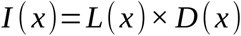

Thus, the present implementation represents an image-normalized directional adaptation of the previously described instability formulation. In the computational workflow, these derived fields were stored as the log-transformed loss-modulus ratio (log_gpp_ratio), the deviation in loss tangent (delta_tand), and the instability field (instability).

For each wedge, mechanical features were summarized from shell voxels on the first post-treatment scan. The principal directional predictors were the mean deviation in loss tangent, the mean log-transformed loss-modulus ratio, the mean instability, and the 95th percentile of instability. Additional exploratory descriptors included the fraction of instability values above predefined thresholds, branch-point density, and skeleton-length density. These secondary features were retained for descriptive analyses but were not the basis of the primary inferential claims.

### Statistical analysis

Descriptive wedge-level analyses related post-treatment mechanical features to later wedge-level novel tumor burden within lesions demonstrating net interval growth. For the continuous outcome, associations were quantified using Spearman rank correlation. For the binary outcome, discrimination was quantified using the area under the receiver operating characteristic curve together with Mann– Whitney comparisons between wedges with and without later novel tumor.

The primary inferential analysis addressed within-patient dependence among wedges by fitting cluster-aware models restricted to lesions with net interval growth. The primary outcome was wedge-level novel tumor fraction. The principal predictors were mean Δtanδ, mean log loss-modulus ratio, mean instability, and the 95th percentile of instability. All predictors were standardized before modelling.

Two complementary modeling strategies were used. Linear mixed-effects models with patient-specific random intercepts were fit to account for correlation among wedges from the same lesion while allowing patient-level differences in baseline novel tumor burden. Generalized estimating equation models were additionally fit with Gaussian family and exchangeable within-patient correlation structure to obtain population-averaged estimates under clustered dependence. Each predictor was modeled separately. Because the wedge-level mechanical features were correlated and the patient-level sample size was limited, multivariable models were not used in order to avoid unstable coefficient partitioning.

Multiplicity adjustment was applied only to the primary inferential family of cluster-aware models. This family comprised the four prespecified mechanical predictors evaluated in one-predictor models for wedge-level novel tumor fraction. False-discovery-rate adjustment was performed using the Benjamini–Hochberg procedure separately within each modeling framework, that is, across the four mixed-effects analyses and across the four generalized estimating equation analyses. Descriptive wedge-level analyses and wedge-width robustness analyses were not included in this correction procedure. Adjusted q values are reported together with nominal p values.

Sensitivity to directional discretization was assessed by repeating the continuous wedge-level association analysis at wedge half-widths of 5, 10, and 15 degrees. For each width, Spearman correlations between the main wedge-level mechanical predictors and later wedge-level novel tumor fraction were recalculated in the growth cohort.

## Results

### Cohort and directional analysis set

Nine patients had analyzable first post-treatment MRE, tumor segmentation at the first post-treatment scan, and second post-treatment MRI suitable for spatial comparison, and were included in the longitudinal post1→post2 dataset (mean age 54, range 25-78). One otherwise eligible patient was excluded because the second post-treatment scan was unavailable. Among the nine longitudinally evaluable patients, six lesions showed net interval growth between the first and second post-treatment scans and were included in the primary wedge-level directional analysis). The remaining three lesions did not show net interval growth and were retained for descriptive comparison only, because they did not provide a positive spatial endpoint for modeling directional tumor emergence. Patient-level clinical and imaging characteristics are summarized in Table 1.

**Table 1:** Cohort characteristics and analysis-set allocation.

| ID | Gender | MGMT promoter status | Pre-treatment volume (cm<sup>3</sup>) | Post-treatment 1 volume (cm<sup>3</sup>) | Post-treatment 2 volume (cm<sup>3</sup>) | Volume change post1 → post2 (%) | Interval post1 → post2 (months) | Included in post1 → post2 analysis | Included in primary growth analysis | Number of 10° wedges |
| --- | --- | --- | --- | --- | --- | --- | --- | --- | --- | --- |
| 1 | Male | Wild-type | 3.8 | 4.68 | 4.56 | -2.7 | 5.98 | Yes | No | 72 |
| 2 | Male | Wild-type | 34.88 | 21.3 | 24.04 | 12.8 | 4.14 | Yes | Yes | 72 |
| 3 | Male | Methylated | 77.71 | 34.57 | 41.59 | 20.3 | 8.8 | Yes | Yes | 72 |
| 5 | Male | Wild-type | 37.54 | 10.17 | 12.13 | 19.3 | 5.62 | Yes | Yes | 72 |
| 6 | Male | Methylated | 32.6 | 33.21 | 38.8 | 16.8 | 15 | Yes | Yes | 72 |
| 7 | Male | Wild-type | 37.04 | 12.54 | 11.21 | -10.6 | 5.35 | Yes | No | 72 |
| 8 | Female | Methylated | 38.71 | 13.74 | 10.73 | -21.9 | 7.1 | Yes | No | 72 |
| 9 | Male | Wild-type | 14.04 | 11.3 | 11.96 | 5.9 | 7.33 | Yes | Yes | 72 |
| 10 | Female | Wild-type | 28.22 | 16.62 | 18.28 | 10 | 11.43 | Yes | Yes | 72 |

At the wedge level, the outcome distribution was sparse, with novel tumor burden concentrated in a minority of directional sectors. This pattern was consistent with the intended design of the framework, which aimed to identify localized directional emergence rather than uniform circumferential expansion.

### Descriptive wedge-level associations

Descriptive wedge-level analyses showed that multiple post-treatment mechanical features were associated with subsequent novel tumor burden in the matched directional wedge (**Table 2**). Among the predictors, the strongest continuous association was observed for the upper-tail instability measure, followed by mean instability and mean Δtanδ (**Figure 2)**. The most consistent binary discrimination was likewise observed for upper-tail instability, with mean instability and mean Δtanδ showing similar but somewhat weaker performance. The mean log loss-modulus ratio showed a smaller positive association.

**Table 2:** Descriptive wedge-level associations between post-treatment mechanical features and later novel tumor burden.

| Predictor | Spearman r with novel tumor fraction | P value (Spearman) | AUC for novel tumor presence | P value (binary) |
| --- | --- | --- | --- | --- |
| Instability, 95th percentile | 2.60E-01 | 4.14E-08 | 7.57E-01 | 4.44E-08 |
| Mean instability | 2.16E-01 | 7.80E-06 | 7.09E-01 | 8.89E-06 |
| Mean $\Delta \tan \delta$ | 1.93E-01 | 6.68E-05 | 6.91E-01 | 4.98E-05 |
| Branch-point density | 1.65E-01 | 7.91E-04 | 6.57E-01 | 8.33E-04 |
| Mean log loss-modulus ratio | 1.02E-01 | 3.70E-02 | 6.00E-01 | 3.00E-02 |

**Figure 2:**
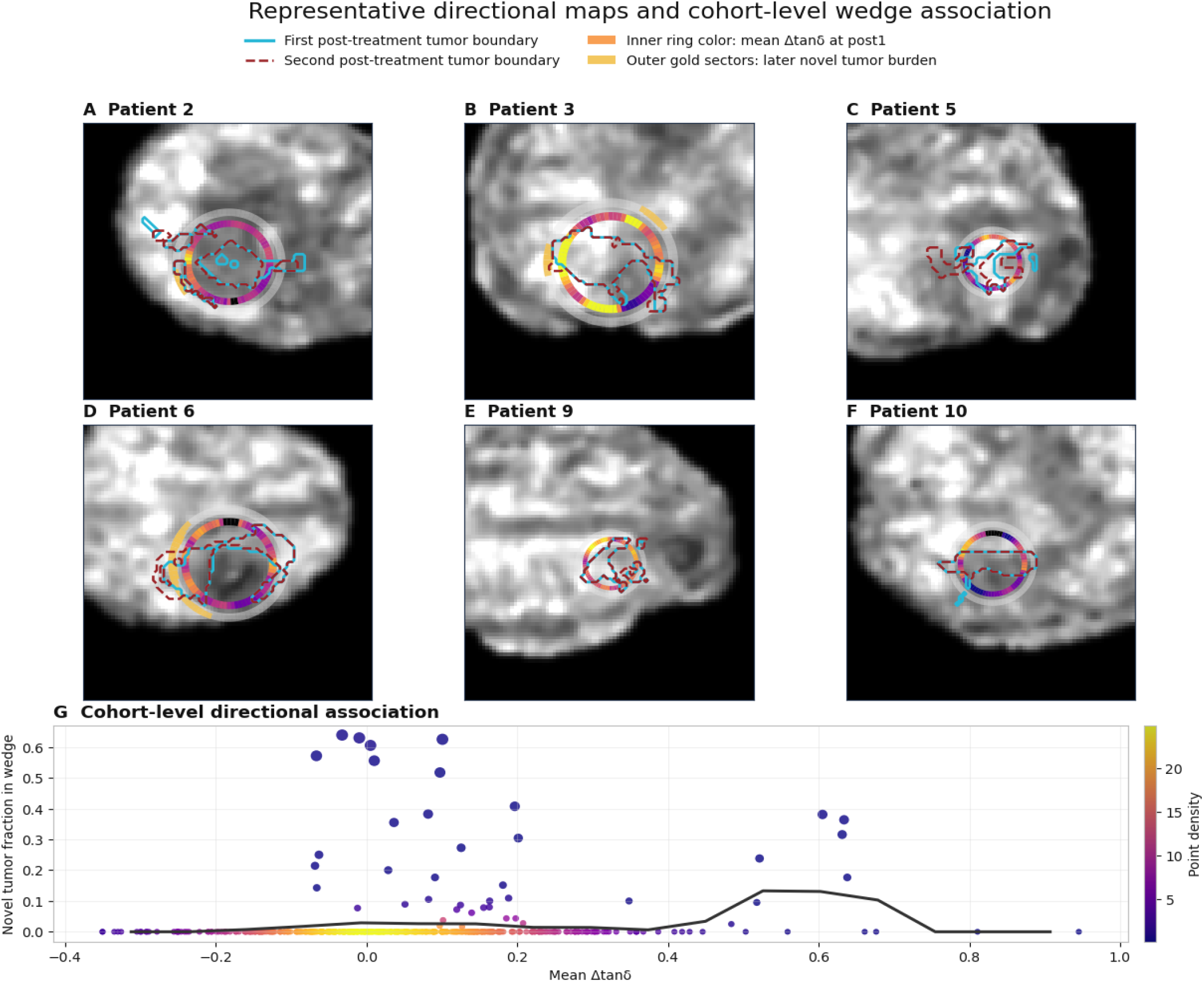
Representative directional interface maps and cohort-level wedge association. Panels A–F show representative lesions included in the longitudinal directional analysis. The cyan contour marks the first post-treatment tumor boundary, and the dashed red contour marks the second post-treatment tumor boundary. Directional sectors were defined around the first post-treatment tumor boundary. The inner ring shows baseline mean Δtanδ at the first post-treatment scan, with warmer colors indicating higher values. The outer gold sectors show later novel tumor burden between the first and second post-treatment scans. Panel G shows the wedge-level cohort association. Each point represents one directional wedge. The x-axis shows baseline mean Δtanδ, and the y-axis shows the fraction of later novel tumor burden in the same wedge. Point color indicates local point density only and is separate from the inner-ring color scale in panels A–F. The black curve shows the smoothed cohort-level trend. Because many wedges had no later tumor emergence, the trend can remain low despite individual high-burden wedges at similar Δtanδ values.

**Figure 3:**
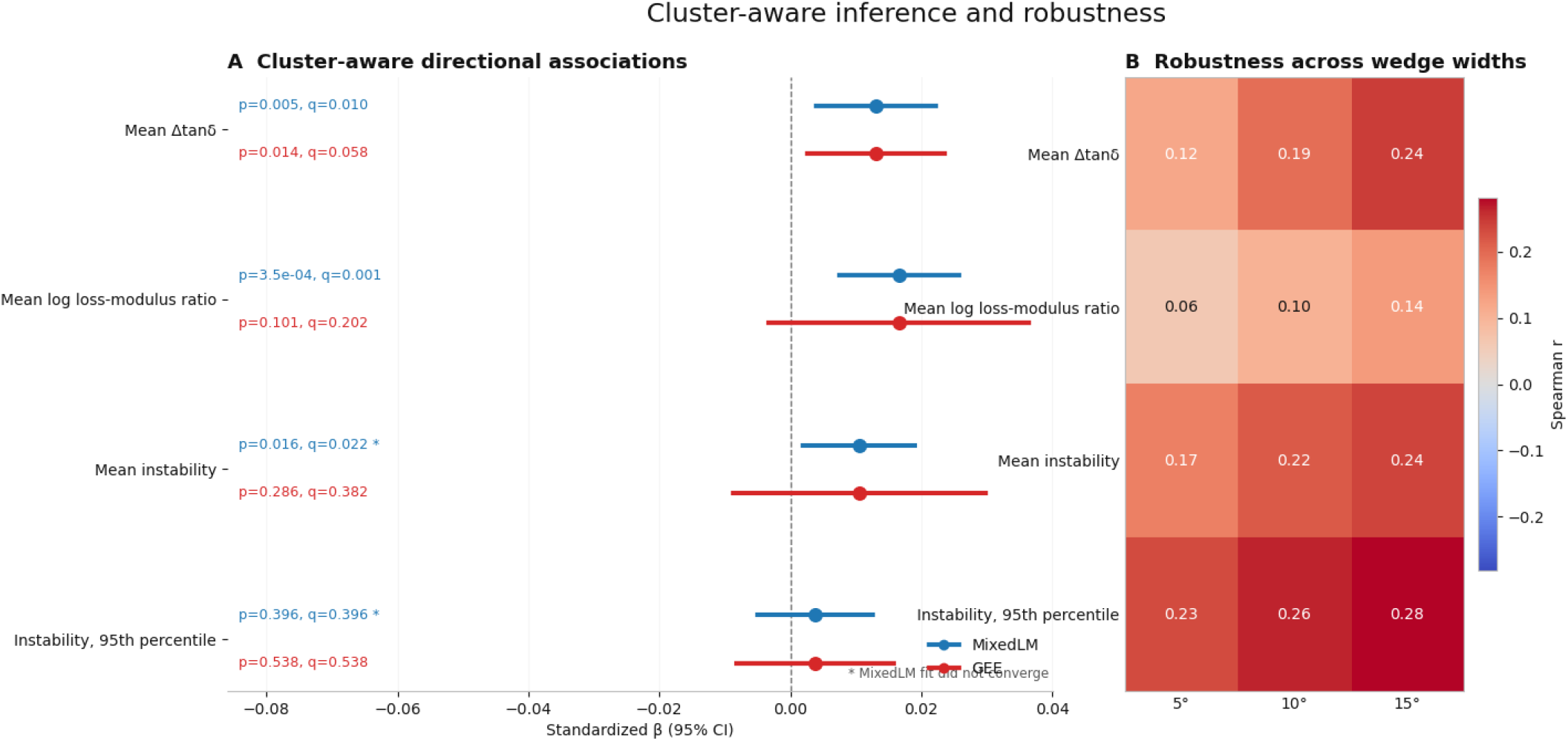
Cluster-aware inference and robustness. Cluster-aware effect estimates for the prespecified directional mechanical predictors of later wedge-level novel tumor fraction are shown for both linear mixed-effects modeling and generalized estimating equations. Points indicate standardized regression coefficients and horizontal lines indicate 95% confidence intervals. Nominal p values and false-discovery-rate adjusted q values are shown for each model; asterisks indicate mixed-effects fits that did not converge. The robustness panel summarizes continuous wedge-level associations between the same predictors and later novel tumor fraction across wedge half-widths of 5°, 10°, and 15° in the growth cohort.

Taken together, these descriptive analyses indicated that wedges with more abnormal post-treatment mechanics tended to show greater subsequent novel tumor burden at the later scan. However, because wedges from the same lesion are not statistically independent, these results were interpreted as descriptive and were not taken as the primary basis for inference.

### Cluster-aware inferential modelling

The primary inferential analysis accounted for within-patient dependence by fitting one-predictor cluster-aware models restricted to lesions with net interval growth (**Table 3**). Across these models, the most robust signal was observed for mean Δtanδ. In the linear mixed-effects model, mean Δtanδ was positively associated with later wedge-level novel tumor fraction (β = 0.013, p = 0.0049, q = 0.0098). This association remained significant in the generalized estimating equation model (β = 0.013, p = 0.014, q = 0.0576), making it the only prespecified predictor that was nominally significant in both modeling frameworks.

**Table 3:** Cluster-aware models of directional mechanical predictors of later novel tumor fraction.

| Predictor | Model | Standardized $\beta$ | SE | P value | FDR q value | Converged |
| --- | --- | --- | --- | --- | --- | --- |
| Mean $\Delta \tan \delta$ | MixedLM | 0.010 | 0.005 | 0.005 | 0.010 | 1 |
| Mean $\Delta \tan \delta$ | GEE | 0.013 | 0.005 | 0.014 | 0.058 | 1 |
| Mean log loss-modulus ratio | MixedLM | 0.017 | 0.005 | 0.000 | 0.001 | 1 |
| Mean log loss-modulus ratio | GEE | 0.017 | 0.010 | 0.101 | 0.202 | 1 |
| Mean instability | MixedLM | 0.011 | 0.004 | 0.016 | 0.022 | 0 |
| Mean instability | GEE | 0.010 | 0.010 | 0.286 | 0.382 | 1 |
| Instability, 95th percentile | MixedLM | 0.004 | 0.004 | 0.396 | 0.396 | 0 |
| Instability, 95th percentile | GEE | 0.004 | 0.006 | 0.538 | 0.538 | 1 |

The mean log loss-modulus ratio also showed a positive association in the mixed-effects model (β = 0.017, p = 3.48 × 10^-4, q = 0.0014), but this effect was attenuated in the population-averaged analysis and was no longer significant in the generalized estimating equation model. Mean instability was similarly associated with later novel tumor fraction in the mixed-effects model (β = 0.011, p = 0.016, q = 0.0216), but not in the generalized estimating equation model. By contrast, the 95th percentile instability measure, despite being the strongest descriptive correlate, was not significant in either cluster-aware framework.

Thus, while several post-treatment mechanical features were descriptively related to later wedge-level tumor progression, the most stable cluster-aware association was observed for the directional phase-shift measure captured by mean Δtanδ (**Table 3**).

### Sensitivity to wedge definition

The direction and relative ranking of the main continuous associations were preserved across wedge widths of 5°, 10°, and 15°, indicating that the findings were not confined to a single arbitrary directional discretization. These sensitivity analyses are provided in **Supplementary Table 1**.

## Discussion

This study shows that local post-treatment interface mechanics in glioblastoma are directionally related to where tumor recurrence occurs. Using a wedge-based longitudinal MRE framework, we found that sectors with greater mechanical abnormality at the first post-treatment scan tended to show more frequent tumor at the second, with mean Δtanδ providing the most robust cluster-aware signal. The main implication is that MRE may capture not only lesion-level biomechanical state, but also spatial information about future growth geometry. This is consistent with prior evidence that glioblastoma exhibits biologically meaningful regional mechanical heterogeneity^6,10,10,24,25,29,32^. Such information could support earlier detection of progression, directional risk stratification, and spatially informed treatment planning such as preoperative radiation and postoperative guiding of local treatment^33,34^.

The principal advance of the present work is the shift from lesion-level description to spatially resolved longitudinal inference. Most imaging biomarkers in glioblastoma indicate whether a lesion has changed, but not where further progression is likely to occur. Here, local mechanics measured at one post-treatment time point were linked to later tumor emergence in matched directional sectors. This supports the view that progression depends on local boundary conditions at the tumor–brain interface^6,10,24,29^.

Among the predictors, mean Δtanδ was the most robust because it remained nominally significant in both cluster-aware frameworks. This suggests that the dissipative phase component of viscoelastic response may be especially informative for identifying interface sectors at risk of later extension. Physically, this pattern is consistent with concepts from rheology and active-matter mechanics in which interfacial instability and finger-like extension emerge when local viscous, elastic and traction-related forces become imbalanced, including phenomena analogous to viscous fingering^8,27,35–38^. Biologically, this signal is consistent with local imbalance between elastic storage and viscous dissipation, as might arise from altered tissue architecture, reduced structural coherence, fluid redistribution, or weakened coupling between tumor and surrounding brain. Viewed in that context, our results support a model in which local mechanical disequilibrium at the tumor-brain interphase, may reflect underlying differences in matrix organization, tissue coupling and stress transmission, thereby biasing later extension toward specific sectors rather than producing uniform bulk growth^12,15,17,25,39–52^.

The behavior of the instability-derived predictors adds nuance to this interpretation. Upper-tail instability and mean instability showed the strongest descriptive relationships with later novel tumor burden, but these associations weakened in cluster-aware models. This suggests that instability measures may be useful as descriptors of local heterogeneity while being less stable as standalone inferential markers in a small clustered dataset. In that respect, the present results complement our earlier cross-sectional instability work: rather than using instability fields primarily for biomechanical phenotyping, the wedge-based framework links local post-treatment mechanics to subsequent directional tumor emergence^6,29^.

The translational relevance of this framework is strengthened by our recent MRE work in post-radiotherapy enhancing lesions, where viscoelastic parameters provided the clearest separation between radiation necrosis and recurrent metastasis and exploratory interface-topology features added complementary spatial information^29^. Together with the current study, this suggests that MRE may be most informative when mechanics are treated as spatially organized rather than reduced to lesion-wide averages. A lesion may show modest change in global summary values while still containing mechanically abnormal sectors that later become sites of novel tumor emergence.

This study has several strengths, including a longitudinal predictor–outcome design, a conservative definition of novel tumor, alignment of the analysis to the tumor-brain interphase, and cluster-aware modeling of within-patient wedge dependence. The main limitations are the modest patient-level sample size, restriction of the primary inferential analysis to growth lesions, use of a maximal two-dimensional slice rather than the full three-dimensional interface, sparse wedge-level outcome structure, and use of an image-normalized adaptation of the previously described instability formulation. The framework was also developed iteratively on the available dataset, and the inferential analyses should therefore be interpreted as exploratory.

Future work should validate this approach in larger prospective longitudinal cohorts, extend it to full three-dimensional surface-based analysis, and determine whether directional mechanical features add predictive value beyond conventional MRI and other advanced imaging markers. The potential of being able to predict the location of glioblastoma recurrence adds to the treatment possibilities including extra focused irradiation therapy and focused opening of the blood brain barrier etc. directed against the areas of highest risk.

## Conclusion

This study provides proof of principle that directional post-treatment MRE can identify biomechanically abnormal interface sectors associated with later tumor emergence in glioblastoma. The findings support a shift from lesion-level mechanical description toward spatially resolved longitudinal prediction and suggest that progression may be shaped by local mechanical conditions at the tumor– brain interface. With validation in larger cohorts, this approach could contribute to earlier directional risk stratification and more spatially informed monitoring of disease evolution. Furthermore, this provides a basic framework for in-vivo studies of brain tumor progression.

## Conflicts of interest

Sandeep K. Ganji is an employee of Philips, Cambridge, MA, United States

## Funding

This work was supported by the Danish Cancer Society (grants R326-A19155 and R352-A20745). Additional support was provided by the National Institutes of Health (NIH) through grants R37-EB001981 and R01-NS113760.

## Data availability

The imaging data and derived wedge-level analysis tables underlying this study are available from the corresponding author upon reasonable request, subject to institutional and patient confidentiality constraints.

## Acknowledgements

We Acknowledge the staff at the Department of Neurosurgery, Odense University Hospital for

---

**Supplementary 1:** Robustness of wedge-level associations across directional wedge widths.

| Predictor | 5° | 10° | 15° |
| --- | --- | --- | --- |
| Mean $\Delta \tan \delta$ | 0.12 | 0.19 | 0.24 |
| Mean log loss-modulus ratio | 0.06 | 0.1 | 0.14 |
| Mean instability | 0.17 | 0.22 | 0.24 |
| Instability, 95th percentile | 0.23 | 0.26 | 0.28 |
| Branch-point density | 0.18 | 0.17 | 0.19 |

## Notes

### Author Declarations

All subjects provided oral and written informed consent prior to inclusion and the study was approved by the Regional Committee on Health Research Ethics for the Region of Southern Denmark (IDs: S-20190105, S-20220055) and was performed according to the declaration of Helsinki.

